# COVID 19: An SEIR model predicting disease progression and healthcare outcomes for Pakistan

**DOI:** 10.1101/2020.05.29.20116517

**Authors:** Ejaz Ahmad Khan, Maida Umar, Maryam Khalid

## Abstract

**Background:** Recent pandemic of the Noval Coronal Virus (COVID 19) has claimed more than 200,000 lives and about 3.8 million infected worldwide. Countries are being gradually exposed to its devastating threat without being properly prepared and with inadequate response. COVID 19’s first two cases were reported in Pakistan on February 26, 2020. We present a model depicting progression of epidemiology curve for Pakistan with and without interventions in view of its health system’ response capacity in near future.

**Methodology:** We used a modified compartmental epidemiological SEIR model to describe the outbreak of COVID-19 in Pakistan including the possibility of asymptomatic infection and presymptomatic transmission. The behavior of the dynamic model is determined by a set of clinical parameters and transmission rate.

**Results:** We estimated that in the absence of a set of proven interventions, the total susceptible population would be 43.24 million, exposed individuals would be almost 32 million, asymptomatic cases would be 13.13 million, mildly infected 30.64 million, severely infected slightly more than 6 million and critical cases would be around 967,000 in number. By that time, almost 760,000 fatalities of infected critical would have taken place. Comparing with the healthcare capacity of Pakistan, if we could “flatten the curve” to a level below the dashed grey line, the healthcare system will be capable of managing the cases with ideal healthcare facilities, where the grey line representing the healthcare capacity of Pakistan. With the intervention in place, the number of symptomatic infected individuals is expected to be almost 20 million.

**Conclusion:** We consider the impact of intervention and control measures on the spread of COVID-19 with 30% reduction in transmission from mild cases in case a set of interventions are judiciously in place to mitigate its impact.

## Background

December 2019 witnessed a new challenge to the advances of Public Health when a novel corona virus emerged on the face of Earth infecting China’s Wuhan province and spread along 219 countries of the World within next four months causing more than four million cases and nearly a quarter of million deaths, mostly in Europe and United States. The COVID 19, as the name given to it, has been identified having three main strains: A, B and C. The ‘B’ strain inflicted Chinese population and mutated to type ‘C’ when it reached Europe after clustering in Singapore(1,2). It also reached Iran and from there to Pakistan. Pakistan had its first report of cases on February 26^th^, 2020(3). Since then, by 16^th^ of May, there have been reported 38,799 cases, with 834 deaths and 10,880 recovered cases(4).

A pandemic need to be understood in the context of its epidemiology and expected number of cases with its case fatality rate and health system’s capacity in dealing with the critical patient load with respect to its surge capacity, in an absence of proper preparedness. We conducted this analysis, based on estimates considering that the cases identified were not the true picture due to insufficient testing capacity and an absence of R_0_ for the country, to model Pakistan’s COVID 19’s trends and the health system’s capacity in dealing with the pandemic. This paper intends to assess the effects of interventions like lock down, social distancing and awareness campaigns in Pakistan via means of reduction in transmission. In the absence of any pharmaceutical aid, the only interventions that can be implemented to avoid the spread of COVID-19 are social distancing, self-quarantine and use of protective gears. The first lockdown was officially imposed on 24^th^ March, 2020 by the government in Pakistan and then eventually moved towards a soft lockdown(4).

## Methods

A modified compartmental epidemiological SEIR model is used to describe the outbreak of COVID-19 in Pakistan including the possibility of asymptomatic infection and presymptomatic transmission. The behavior of the dynamic model is determined by a set of clinical parameters and transmission rate(5,6).The classic SEIR model assumes that an individual in the population is susceptible (S), exposed (E) where they are asymptomatic and do not transmit infection, infected and infectious (I), or recovered and immune (R) (7,8). It is assumed that when an individual is symptomatic and infected, the progression from exposed stage (E) to becoming infected and infectious (I) occur at the rate *α*. The model considers three clinical stages of infections: mild, severe and critical (9) (refer to as *I*_1_, *I*_2_, *I*_3_ onwards) (10). Each infection stage of an individual has two possible outcomes: recovery and progression towards next stage leading to death. Let *γ*_1_, *γ*_2_, *γ*_3_ be the recovery rates at each of the three infectious stages respectively and let *ρ*_1_, *ρ*_2_ be the progression rates from *I*_1_ to *I*_2_ and from *I*_2_ to *I*_3_ respectively. While let *δ* be the transition rate of the death (D) an individual at *I*_3_. Let *θ*_1_, *θ*_2_, *θ*_3_ be the respective transmission rates at the three stages *I*_1_, *I*_2_, *I*_3_ of the model.

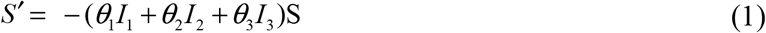

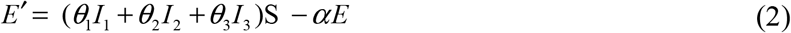

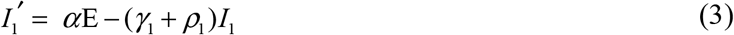

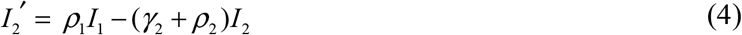

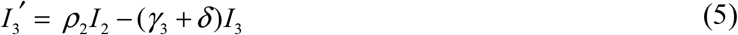

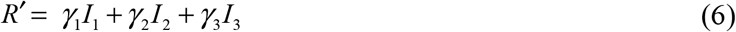

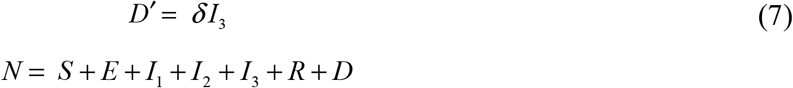

where

*θ*_1_ = the rate at which the infected individual in class *I_i_* contact susceptibles and infect them

*α*_1_ the rate of progression from exposed to infected

γ_i_ = the rate at which the infected individual in class *I_i_* recover from the disease

*ρ_i_* = the rate at which the infected individual in class *I_i_* progress to the next class I_i+1_

δ = the death rate of individuals in most severe stage of infection.

*S*: Susceptible Individuals

*E*: Exposed Individuals (Infected but asymptomatic)

*I_i_*: Infected Individuals in class *i* for *i* = 1, 2, 3

*R*: Individuals Recovered and are now immune

*D*: Individuals died of infection

*N*: Total population

The parameters of the model define above are then estimated as follows, incorporating the information from clinical data.

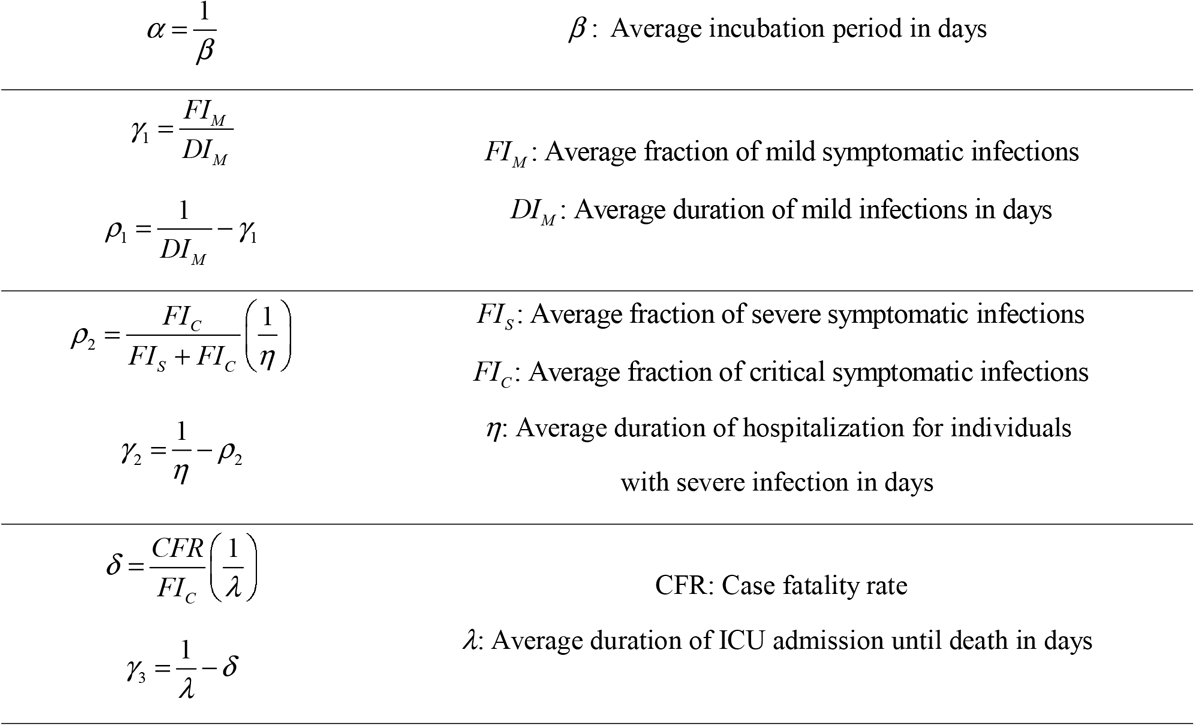

Let *r* be the exponential growth rate for the epidemic, at initial stage, with the doubling time 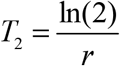. The ratio of individuals during the initial stage of any epidemic is fixed between any two compartments and their expected values can be computed using the underlying model with fixed parameters. The expected ratios are given as follows:

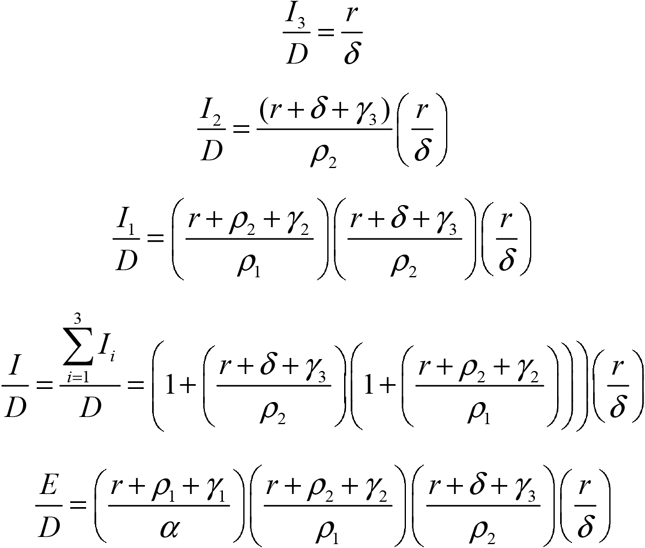

These expected ratios can further be used to determine the number of under reported cases under the following assumptions:

i. The model is formulated as a system of differential equations and its output represents the expected values of each quantity. Further, as the stochastic events are not considered therefore it cannot be assumed that the epidemic will extinct even for the very low values Also, the model does not report the expected variance in the variables, which can sometimes be large.
ii. All individuals have equal transmission rates and equal susceptibility to infection.
iii. Individuals must pass through stage before reaching stage and only the individuals at critical stage die.

The simulation study assumes *N* = 220,892,340 and *Initial Infected* = 2 for Pakistan. The parametric values are either adapted from the literature or from the official statistics (Table 1) and (Table 2).

**Table 1:** Estimated parameters for COVID-19 clinical progression.

| Parameter | Value |
| --- | --- |
| $\beta$ | 5 days |
| $FI_M$ | 80% |
| $FI_S$ | 15% |
| $FI_C$ | 5% |
| $DI_M$ | 6 days |
| $DI_S$ | 8 days |
| $DI_C$ | 8 days |
| $\eta$ | 14 days |
| $\delta$ | 72% |
| CFR | 2% |
| Time from symptoms to ICU admission | 12 days |
| Time from symptom onset to death | 20 days |
| Serial interval | 8 days |

**Table 2:** Transmission rates for COVID-19.

| Quantity | Value |
| --- | --- |
| $\theta_0$ | 0.5/day |
| $\theta_1$ | 0.27/day |
| $\theta_2$ | 0.1/day |
| $\theta_3$ | 0.1/day |
| $\theta_E$ | 0.5/day |
| $\pi$ | 30% |
| $\mu$ | 6 days |
| $R_0$ | 3.2 |
| $r$ | 0.17/day |
| $(\beta - \beta_1)$ | 2 days |

Despite close monitoring, there are chances of asymptomatic COVID-19 infection (11). This paper incorporates the possibility of such infections by assuming that all the individuals progressing from E to I can be categorized as either asymptomatic or symptomatic. Let *π* be the proportion of individuals with asymptomatic infections such that they progress to *I*_0_ from the stage E, whereas (1 − *π*) be the proportion of symptomatic individuals that progress to *I*_1_ from the stage E. Thus the asymptomatic exposed class can be categorized into two sub-classes namely *E*_0_ (no symptoms and no transmission) and *E*1 (no symptoms but transmit at the rate *θE*) with the rate of exit being *α*0 & *α*1 from each sub class respectively. It is pertinent to note that *E*_1_ represents the pre symptomatic class where delayed symptoms are observed relative to when an individual actually got infected with COVID-19.

Assuming that the asymptomatic infections cannot progress to severe stages and have a transmission rate *θ*0, let *γ*_0_ be the recovery rate of such individuals. Furthermore, all the transmission rates are modified to incorporate the seasonality factor *σ* (t) = 1 + *ɛ* cos(2*π* (t-*φ*)) where *ɛ* ∈[0,1] is the relative amplitude of seasonal oscillation and *φ* ∈[−∞, ∞] is the phase that determine the peak time (in years) relative to starting time. The customized SEIR model is thus given as:

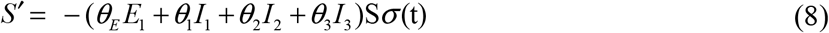

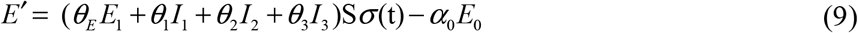

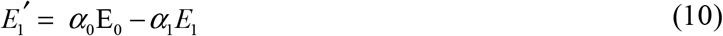

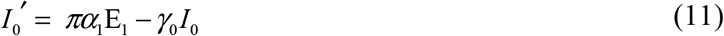

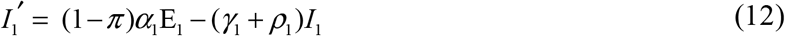

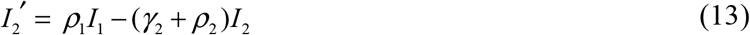

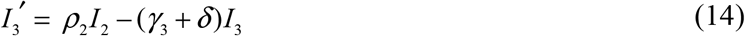

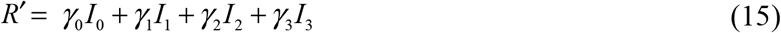

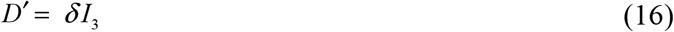

where

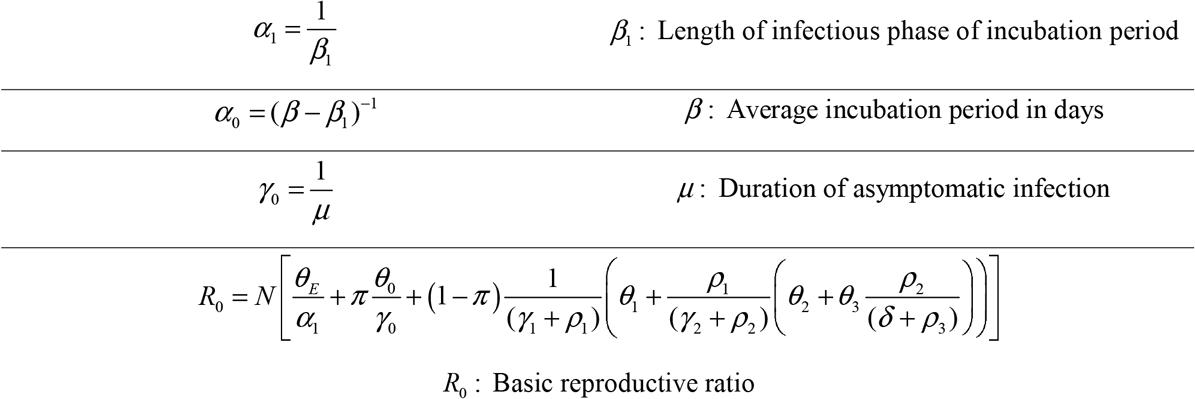

We considered the impact of intervention and control measures on the spread of COVID-19 with a 30% reduction in transmission from mild cases. (12–14).

Given the intensity of the COVID-19 pandemic, where the most developed countries are falling short in allocating appropriate medical facilities to all COVID-19 patients, an under-developed country such as Pakistan has to be very cautious and concerned about optimizing its almost non-existent healthcare facilities relative to its population size. In this regard, it becomes important to analyze the health care capacity along with the COVID-19 estimates and predictions. The analysis is based on the indicators such as hospital beds (per capita), ICU beds (per capita) and ventilators taking into account the effect of intervention to assess the shortfall of resources (Table 3)

**Table 3:** Hospital capacity indicators.

| Quantity | Total | Per capita | Source |
| --- | --- | --- | --- |
| Hospital beds | 132227<sup>a</sup> | 0.60 | (Pakistan Bureau of Statistics, 2018) |
| Occupancy | 80% |  | (Adam T et al, 2003) |
| ICU beds | 3142 | 0.01 | (Phua J et al, 2020) |
| Occupancy | 80% |  | (Adam T et al, 2003) |
| Ventilators | 3800 | 0.02 | (Pakistan Institute of Development Economics, 2020) |

## Results

### Predicted COVID-19 cases of Pakistan by clinical outcome

Figure 1 shows the progression of the epidemic in Pakistan, displaying the changes in the expected number of individuals who are susceptible, exposed, infected and removed as per the modified SEIR model. As the infected cases first experience an asymptomatic phase (incubation period) followed by an infected phase that varies in intensity, the Infected individuals have been further categorized into infected-asymptomatic; infected-mild (I_1_); infected-severe (I_2_) and infected-critical (I_3_) whereas the Removed category has been sub-divided into recovered and dead depending upon the end result of the infection. Using the R_0_ = 3.2 and r = 0.17, it is predicted that the inflection point/peak is expected to fall on the 117^th^ day (21^st^ June, 2020) in Pakistan. The rate at which the infection would be spreading according to this model is reflected by T_2_, which would be 4.1 days. The total susceptible population would be 43.24 million, exposed individuals would be almost 32 million, asymptomatic cases would be 13.13 million, mildly infected 30.64 million, severely infected slightly more than 6 million and critical cases would be around 967,000 in number. By that time, almost 760,000 fatalities of infected critical would have taken place.

**Fig 1:**
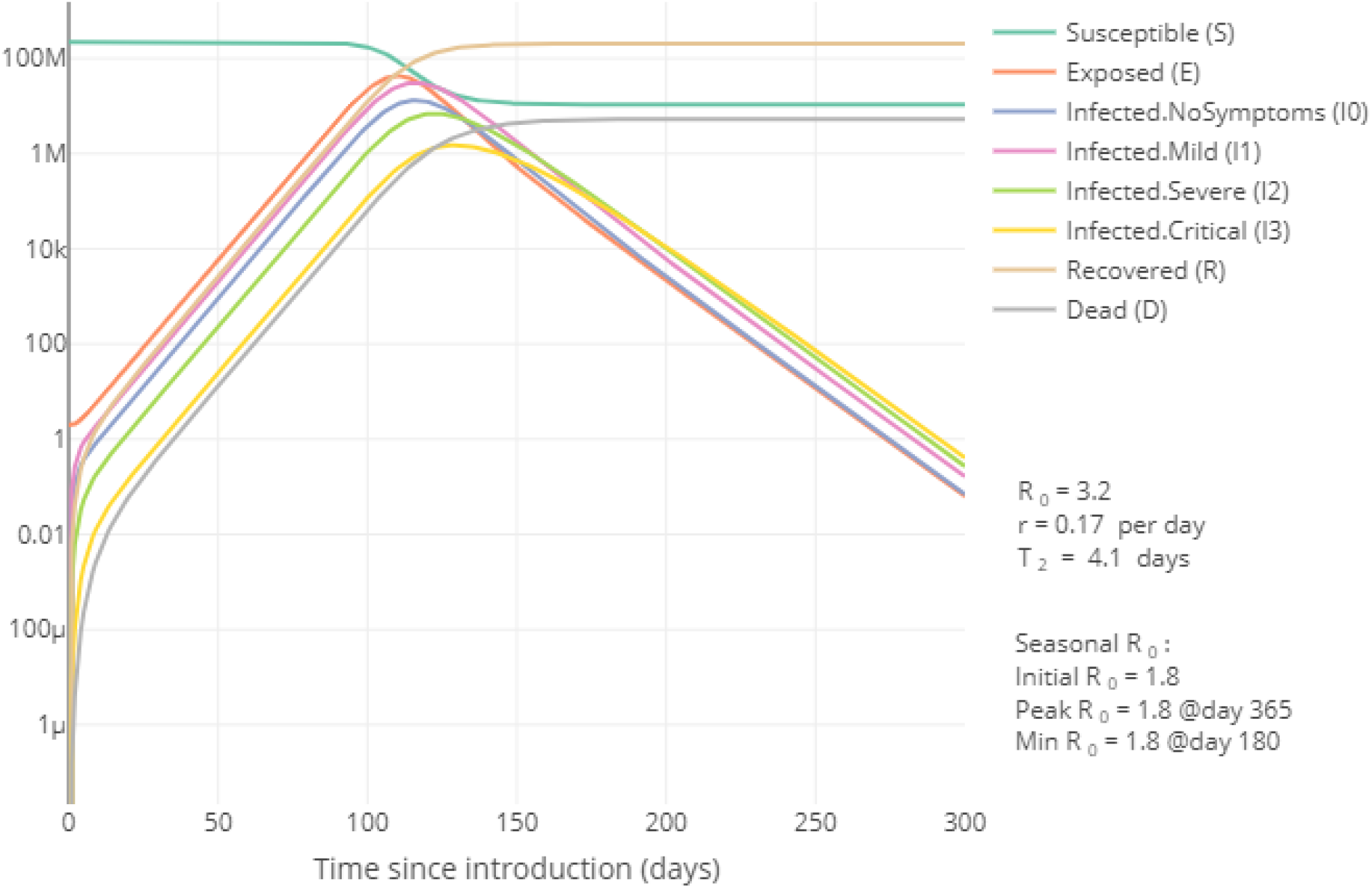
Predicted COVID-19 cases of Pakistan by clinical outcome

### Reduction in predicted COVID-19 infected cases of Pakistan after intervention

Figure 2 shows the changes in the expected numbers of individuals who are symptomatic irrespective of the severity of the infection with and without an intervention being implemented to control the spread of the infection. With the intervention in place, the numbers of symptomatic infected individuals (I_1_+I_2_+I_3_) are expected to be almost 20 million while without intervention; this number rises to a maximum of almost 38 million at the peak of the epidemic. Note the change in R_0_ as it moves from 3.174 without the intervention to a value of 2.8338 with any intervention. Also noteworthy is the change in doubling time, which increases to 5 days upon implementation of intervention instead of 4 days without intervention depicting the accelerated spread without any control measures in place.

**Fig 2:**
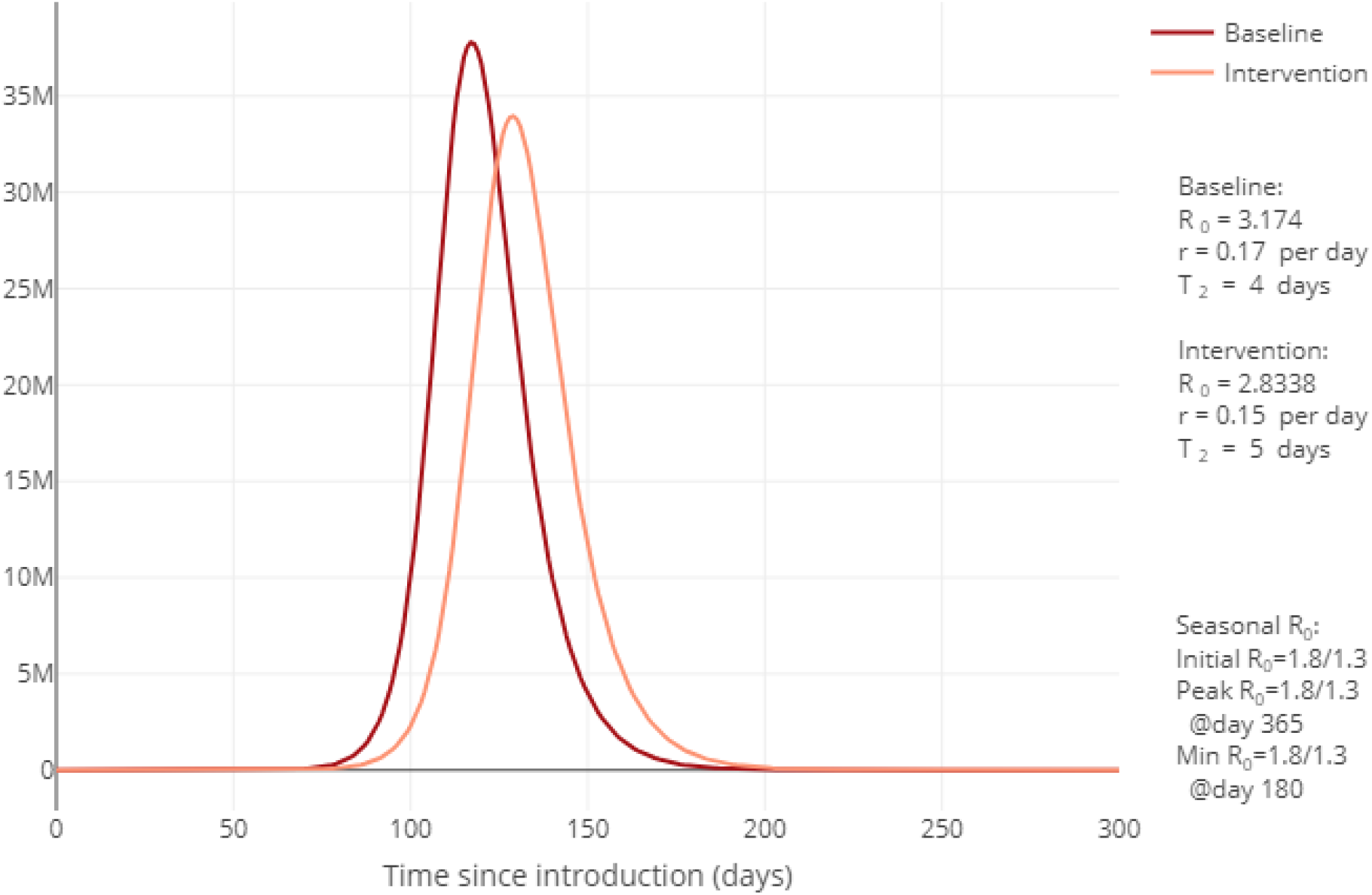
Reduction in predicted COVID-19 infected cases of Pakistan after intervention

### Reduction in predicted COVID-19 deaths of Pakistan after intervention

Almost 5 million deaths could be expected without intervention keeping the Case Fatality Rate (CFR) of Iran of 3.6 as the standard, which can be reduced to 4.39 million deaths if interventions are being implemented to control the spread.

**Fig 3:**
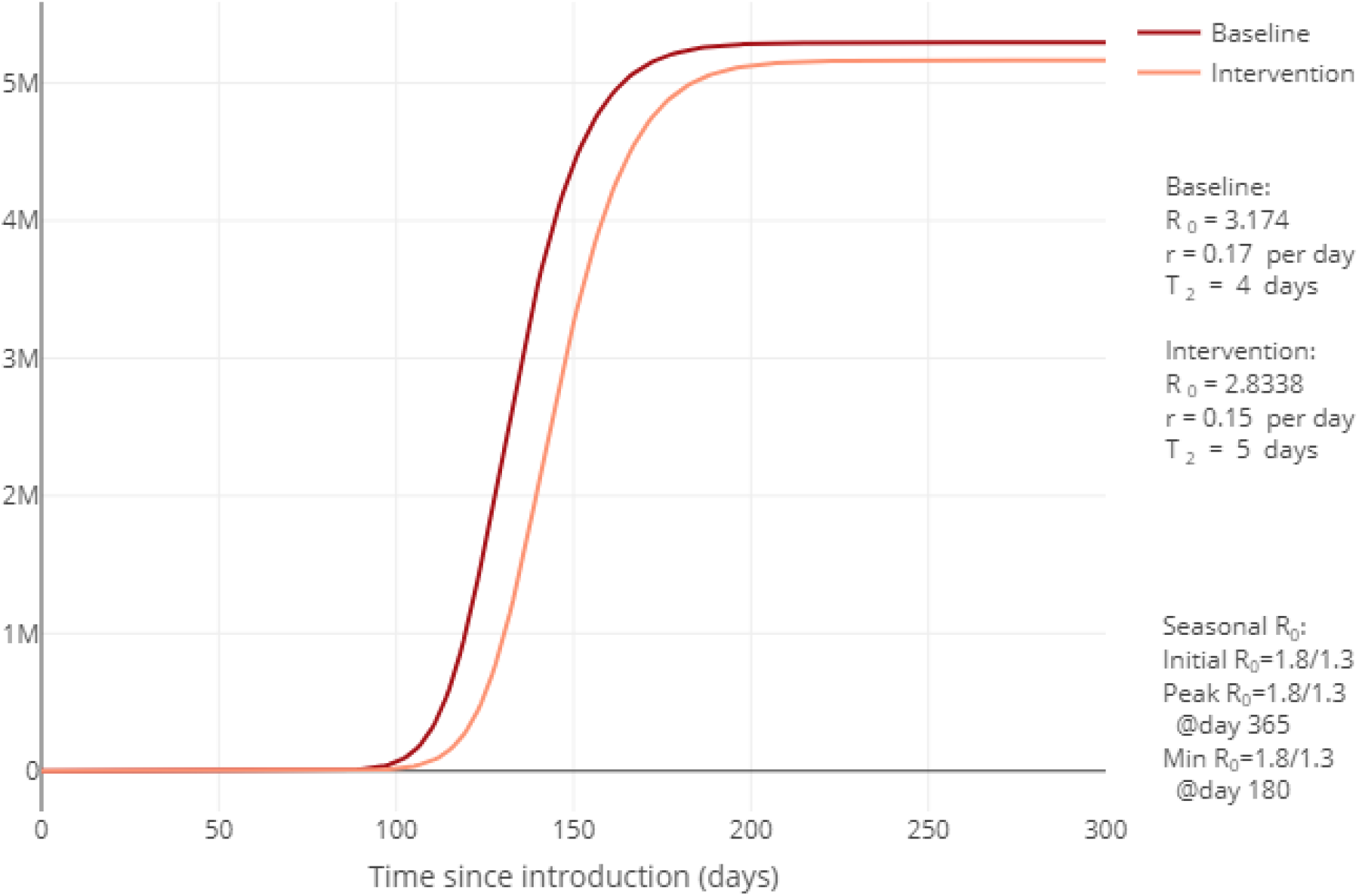
Reduction in predicted COVID-19 deaths of Pakistan after intervention

### COVID-19 cases versus healthcare capacity of hospital beds in Pakistan

Figure 4 shows the number of COVID 19 infections in comparison with the healthcare capacity of Pakistan. If we can flatten the curve to a level below the dashed grey line, the healthcare system will be capable of managing the cases with ideal healthcare facilities, the grey line representing the healthcare capacity of Pakistan.

**Fig 4:**
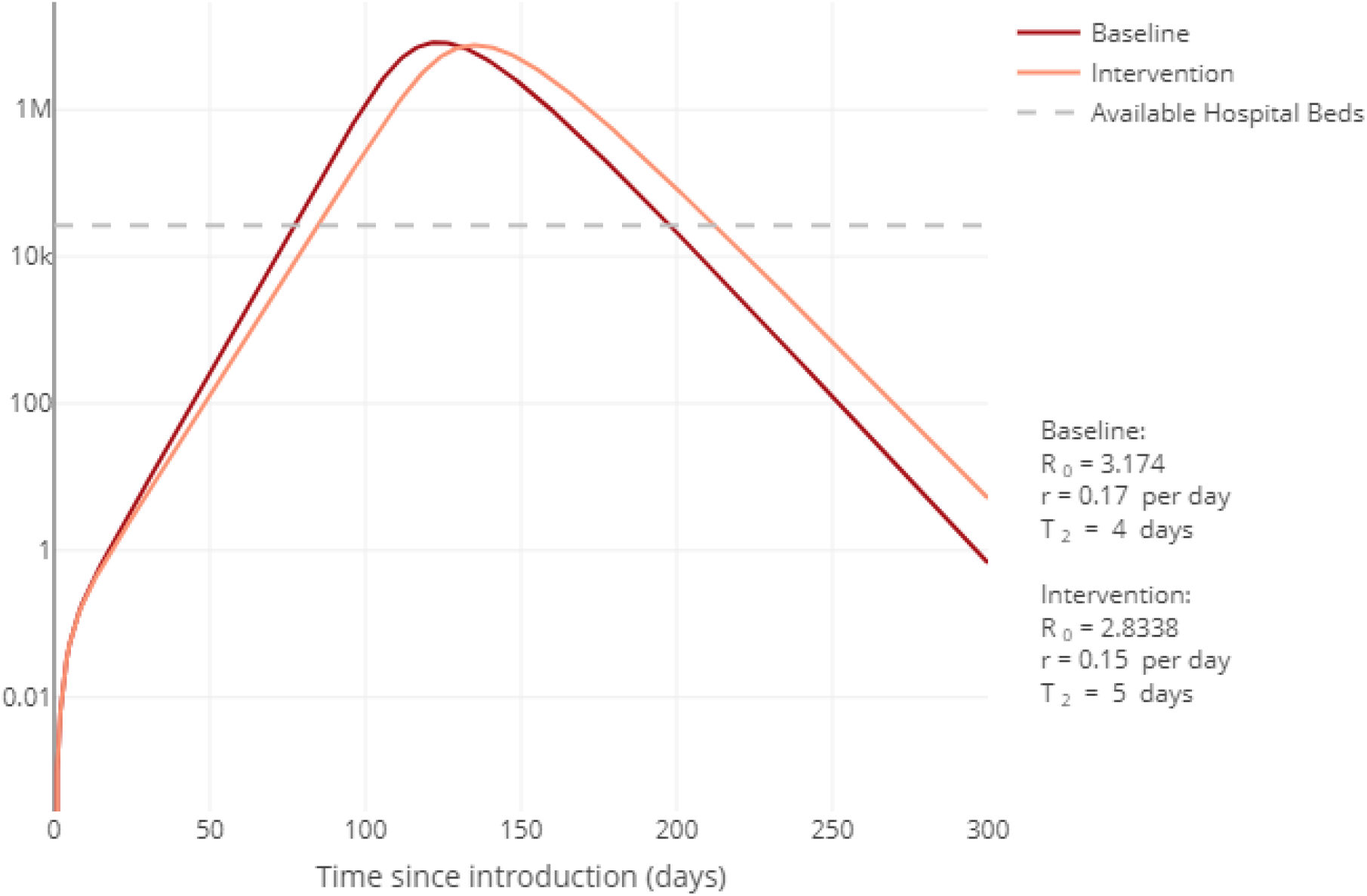
COVID-19 cases versus healthcare capacity of hospital beds in Pakistan

More than seven million individuals with critical and severe infections would need hospitalization while hospital beds are available for only 26,500 individuals at present.

### COVID-19 cases vs healthcare capacity of ICU beds in Pakistan

More than 1 million patients would need ventilation and intensive care and considering the number of ICU beds available, the current available capacity of ICUs is around 0.01 per capita to cater to the needs of one million patients.

**Fig 5:**
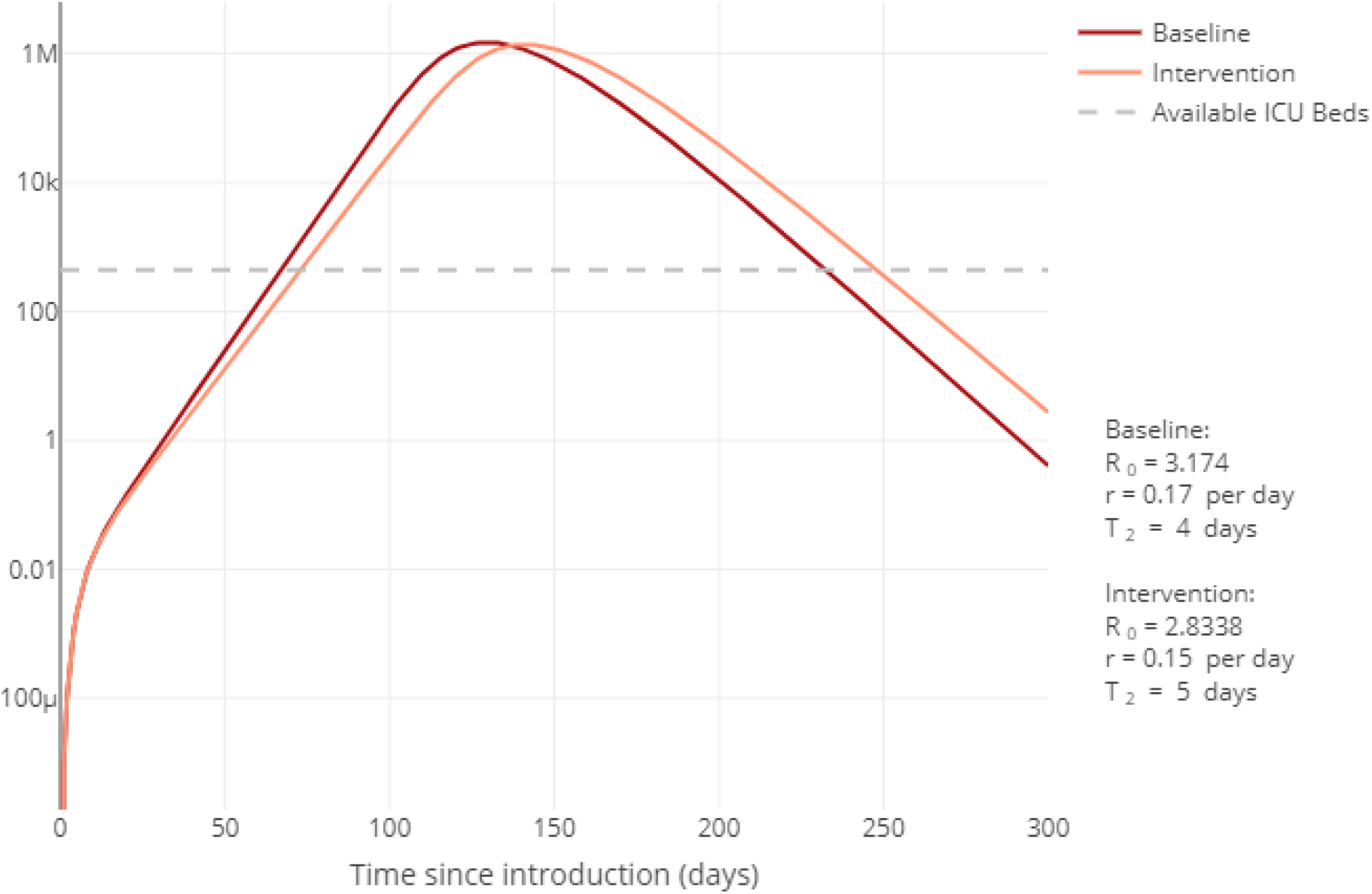
COVID-19 cases vs healthcare capacity of ICU beds in Pakistan

### COVID-19 cases vs healthcare capacity of ventilators in Pakistan

More than 1 million patients would need ventilators along with intensive care and considering the current number of ventilators, only 0.02 per capita would be available.

**Fig 6:**
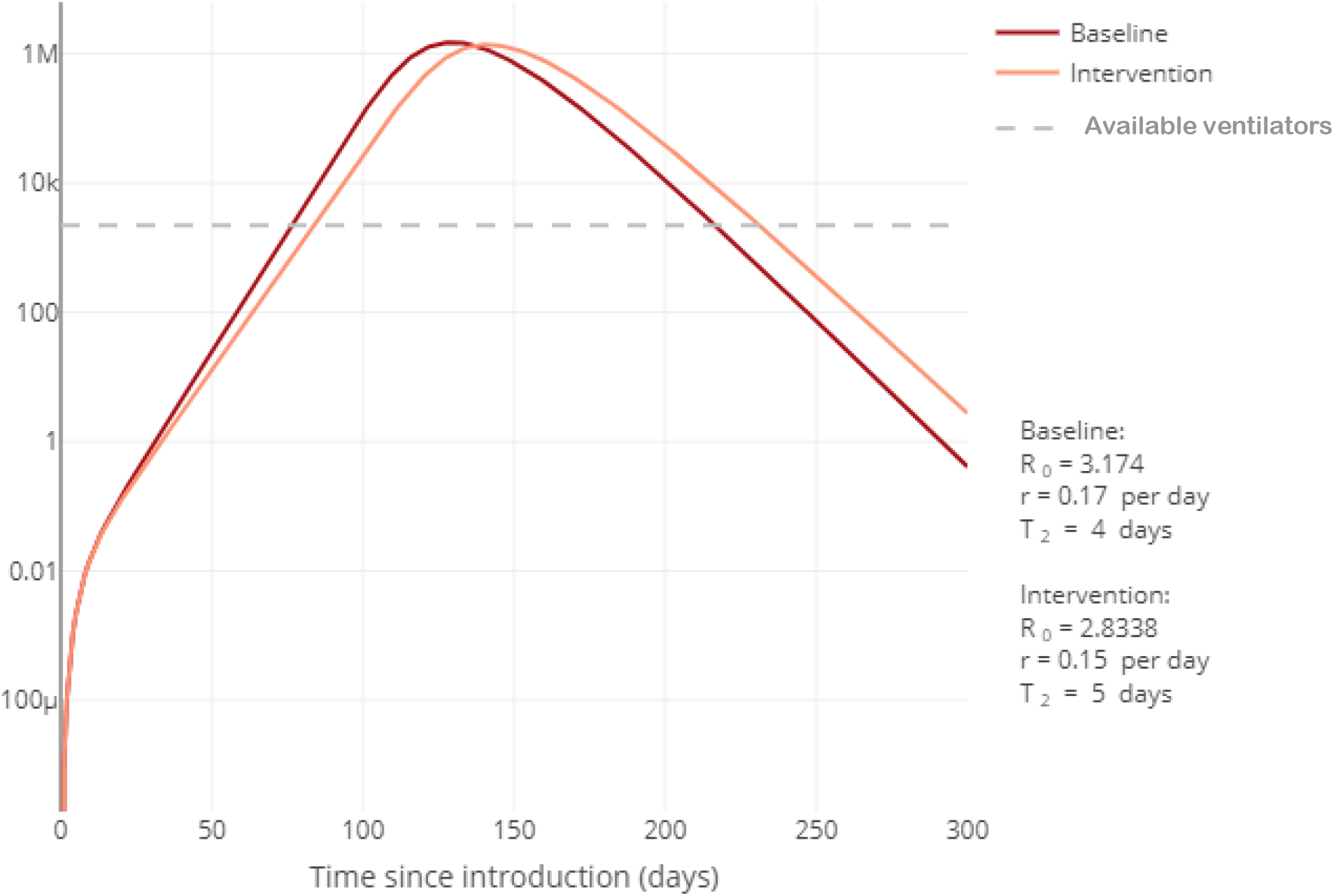
COVID-19 cases vs healthcare capacity of ventilators in Pakistan

## Discussion

Our results show how the susceptible cases would increase over time along with expected number of deaths. These figures look a bit exaggerated when compared to the reported data. But they have similar results and trends for this pandemic done for other countries currently experiencing some of the highest numbers of cases and deaths. As the recovered increase, the susceptible population goes down. The country’s peak is however yet to be seen(15–17). Our results of with and without intervention show a small impact of an intervention (nearly 10 million less cases) as compared to number of expected cases, in reducing transmission of diseases with “r” = 0.17 dropping to r = “r” = 0.15 with intervention. This small reduction with a single intervention has been well documented (18). Timely interventions could reduce the expected cases almost by half, and the expected load on the health system’s capacity, which is quite limited. This could be possible only by introducing appropriate interventions for reducing community transmission by social distancing, identifying cases and contacts by sufficient number of tests for isolating the contacts/suspected cases in quarantines and treatment of cases under suitable infection control conditions taken care of by healthcare personnel protected by personal protective equipment (PPE). The severe infected cases would need hospitalization while those in critical condition would require ventilators and care in Intensive Care Units (ICUs)(19–23). Flattening of the curve and bringing it down with recommended interventions could not only compensate for the limited capacity of the ICUs’s capacity and need for ventilators, it would also improve the efficiency and preparedness of the system in saving lives(18,24,25).

## Conclusion and Recommendations

We conclude that Pakistan’s COVID 19 outbreak is yet to see its see its peak probably towards the third week of June 2020. The cases and admissions in emergency and critical care would compromise the working of the health system. Timely implementation of the prudent interventions would minimize the morbidity and mortality due to the pandemic and would support the weak health system in responding to the outbreak. Pakistan need to invest in the resilient health system and robust pandemic preparedness in place to address control measures for a pandemic as large and devastating as COVID 19.

## Limitations and strengths

Our results are based on assumptions and subsequent predictions based on modeling. These may not be accurate. The mathematical models can guide and inform of a magnitude of disease or event. It is imperative that the policy makers may make their own informed judgement in the best interest of providing timely preventive and curative healthcare. The study’s strength lies in its methodical approach, which is the best available SEIR model available to date for predicting the course of a pandemic.

## Data Availability

All data is available in the manuscript and attached figures.

## Acknowledgements

We acknowledge work done and published by different renowned scientists, whom we have cited in our article. Without this knowledge base, we would not have been able to make these scientific predictions.

## Conflict of interest

We declare that we do not have any conflict of interest whatsoever in conducting this analysis and drafting the manuscript.

### Authors’ contribution

Ejaz Ahmad Khan conceived the idea, design of the study and helped in analysis, and finalized the manuscript.

Maida Umar conducted the analysis wrote the results and helped in writing methods.

Maryam Khalid wrote the method section.

## Notes

### Competing Interest Statement

The authors have declared no competing interest.

### Clinical Trial

Our study was a modeling exercise and NOT a clinical Trial

### Funding Statement

WE declare that we received no funding whatsoever for conducting this study

### Author Declarations

The IRB approval was not required as the study used secondary published data and assumptions for modelling purposes.

